# Blood pressure following resistance exercise in individuals in the chronic phase of stroke recovery

**DOI:** 10.64898/2026.09.18.26363411

**Authors:** Juliano A Abreu, Elric Y Allison, Kenneth S Noguchi, Baraa Al-Khazraji, Ada Tang

## Abstract

Current evidence supports resistance training for stroke recovery but autonomic dysregulation following stroke can lead to exaggerated fluctuations in blood pressure (BP) in response to exercise, thus challenging perceptions about safety and feasibility of resistance exercise. This study compared the post-exercise BP response following a single bout of leg-press exercise between people with and without stroke. Data from 11 individuals with chronic stroke (age 39-76 years, 5 female, 1-9 years post-stroke) and 19 recreationally active controls (age 18-33 years, 11 female) were included. Brachial artery BP was measured before exercise and immediately following a single set of leg-press (8 repetitions, target rating of perceived exertion 5-7/10). The difference between pre- and post-exercise BP was calculated and compared between groups using an independent t-test of equal variance. Additionally, a two-way analysis of covariance was used to examine potential cofounding effects of activity level and baseline BP on between group differences in BP response. No group differences were observed in post-exercise change in systolic (mean difference=3.61 mmHg, 95%CI:-8.36, 15.57, p=0.54); and diastolic (mean difference=5.38 mmHg, 95%CI:-6.11, 16.86, p=0.35) BP. Results were unchanged after controlling for baseline BP, and activity levels. Our work suggests that individuals with and without chronic stroke experience similar acute BP responses immediately following resistance exercise; however, work with more appropriately matched controls and greater sample sizes is needed to support these findings. Future studies may elucidate mechanisms related to the acute cardiovascular response to resistance exercise in individuals with stroke.

## INTRODUCTION

Stroke is the leading cause of adult neurological disability where approximately 63% of people with stroke experience ongoing disability.^1^ Physical consequences from stroke include muscle weakness,^2,3^ which contribute to inactivity^4^ and further declines in health and quality of life.^1,5^

Although current guidelines for stroke recovery recommend a combination of aerobic and strength training activities,^6,7^ most of the literature in stroke rehabilitation focuses on aerobic training to improve cardiovascular fitness, endurance, cognition, and recurrent stroke.^4,8,9^ More recently, resistance training has shown to improve other aspects of life deemed important to people living with stroke, such as lower body strength, walking speed, mobility, balance and activities of daily living;^10,11^ however, concerns may exist among clinicians regarding the blood pressure (BP) response to resistance exercise in people with stroke.^6,12^

Autonomic dysfunction is often impaired following stroke, impacting an estimated 25-76% of individuals,^13–15^ commonly resulting in delayed BP regulation^16^ and increased BP variability which may persist into the chronic stages of stroke recovery.^15,17^ During exercise, while BP increases due to increases in cardiac output and vascular resistance,^18^ post-exercise hypotension may occur^19,20^ when the relaxation of previously active skeletal muscles weakens the muscle pump effect,^21^ temporarily decreasing cardiac output.^21^ Additionally, vasodilators, such as prostaglandin II and nitric oxide, are released during exercise and can remain active for extended periods of time,^22^ contributing to prolonged vasodilation,^23^ decreasing peripheral vascular resistance following exercise.^22^ In individuals with stroke who exhibit autonomic dysfunction and delayed cardiovascular responses to intense exercise,^24^ rapid fluctuations in BP following exercise may increase risk of faints and falls.^25^

Prior work on the post-exercise BP response after stroke has demonstrated significant reductions in BP following aerobic exercise (−4.7mmHg to -11mmHg),^26^ but this relationship has not yet been examined following acute bouts of resistance exercise in this population. Given that resistance exercise typically induces larger fluctuations in BP^27^ compared to aerobic exercise due to the mechanical compression of blood vessels from contracted muscles, the cessation of more intense muscular contractions following acute resistance exercise may lead to greater reductions in BP following acute resistance exercise compared to aerobic exercise.^28^ Due to the substantial differences between acute resistance and aerobic exercise stimuli, there is a need to better understand the BP response following acute resistance exercise in individuals living with stroke.

The purpose of this study was to determine if individuals with stroke exhibit a greater reduction in systolic BP (SBP) and diastolic BP (DBP) immediately following a single bout of lower body resistance exercise compared to individuals without stroke. We hypothesized that individuals with stroke will exhibit a greater decrease in SBP and DBP following a single bout of resistance exercise compared to individuals without stroke, consistent with the literature in similar populations with autonomic dysfunction (i.e., hypertension) following resistance exercise.^29,30^

This study will provide insight into the extent of BP response to acute resistance exercise and thus help guide exercise prescription in stroke populations.

## METHODS

This was an analysis of data from two trials in people with and without stroke. Both trials were approved by the Hamilton Integrated Research Ethics Board (stroke #16189, non-stroke comparison group #16441) and participants provided informed and written consent to participate. The first is a secondary analysis of exercise data from a clinical trial in people with stroke (ClinicalTrials.gov ID: NCT05816811),^31^ and second trial is a prospective intervention study designed to match the same acute resistance exercise stimuli and timing of measurements.

### Experimental Procedures and Protocols

Data for the stroke group were collected as part of a 10-week resistance training intervention for individuals with stroke.^31^ In brief, this was a single-group feasibility trial in people with mild to moderate chronic stroke. This resistance exercise was performed in a community gym and post-exercise BP was collected following every exercise set.^31^ For the purpose of the current study, exercise data from the first set of the horizontal leg press exercise (8 repetitions, Technogym Selection 900 Leg Press, Mississauga, Canada) in Week 2 were used. This exercise set was selected as it was the first exercise of the day to limit the effect of prior resistance exercise on BP. Target intensity was a rating of perceived exertion of 7/10 on the modified Borg scale.^32^ Brachial BP was measured (Omron IntelliSense Professional Digital Blood Pressure Monitor (model HEM-907XL), Kyoto, Japan) as discrete single measurements to reflect common practice in community exercise programs before and within 1 minute following the exercise set, given that the rate of BP decline after exercise is largest in the immediate 30 seconds.^33,34^

Data for non-stroke controls were collected from a study examining physiological responses to acute resistance exercise in healthy, recreationally active-young adults. For the current analysis, the exercise stimulus (intensity and exercise (leg press)) and timing of BP measurements of the non-stroke group were designed prospectively to be similar to that of the stroke group. Thus, we used data from the first set of 8 repetitions of semi-recumbent leg press at 50% 1 repetition max to closely approximate the intensities achieved by the stroke group (5/10 modified Borg Scale ≈ 50% 1-RM).^35,36^ Brachial artery BP was measured (Omron IntelliSense Professional Digital Blood Pressure Monitor (model HEM-907XL), Kyoto, Japan) before and within 1-minute following the exercise set to match methods for the stroke sample. Rating of perceived exertion was also collected for comparison of exerted intensity with the stroke group.

To compare weekly activity levels between groups, the World Health Organization Global Physical Activity Questionnaire was administered to both groups, which has demonstrated adequate reliability and validity in assessing levels of moderate to vigorous physical activity in individuals with^37^ and without^38^ stroke.

### Participants

Individuals with stroke were included if they were: i) ≥18 years old, ii) ≥6 months post-stroke,

iii) able to walk >10 meters with or without the use of an assistive device, iv) living in the community, and v) had mild to moderate stroke severity (modified Rankin Scale ≤3). Individuals were excluded if they: i) had any contraindications to exercise for people with cardiovascular disease according to the American College of Sports Medicine, such as unstable angina, uncontrolled hypertension, orthostatic BP with exercise, or uncontrolled arrhythmias^39^; ii) had significant cognitive impairment (Montreal Cognitive Assessment Blind score <18) that would preclude safe exercise participation; iii) were actively engaged in stroke rehabilitation services, defined as a personalized exercise program for a set duration of time, which is supervised and prescribed by a rehabilitation professional.

The non-stroke group included untrained individuals without stroke who were ≥18 years old. Individuals were excluded if they: i) were pregnant; ii) had a mild traumatic brain injury in the last year; iii) had any history of severe traumatic brain injury, cardiovascular, cerebrovascular or neurological conditions, metabolic syndrome, syncope or light-headedness, migraines or chronic headache, respiratory illnesses, diabetes, claustrophobia; iv) had any current musculoskeletal injury at time of study that would make it difficult or unsafe to perform resistance exercise; v) had been taking any anabolic steroids or any other banned performance enhancing substances as outlined by the Canadian Centre for Ethics in Sport.^40^ Age matching was not applied in the current study due to convenience sampling at a university setting resulting in the recruitment of predominantly younger adults.

### Statistical Analysis

Descriptive statistics were reported using mean (standard deviation) or median [interquartile range] for normally and non-normally distributed data, respectively, and as frequencies (n, %) for categorical data. A scatter plot was used to visually inspect the data. Normality was assessed visually using a histogram and using a Shapiro-Wilks test. Levene’s test was used to assess the variance between groups. For the unadjusted analysis, two independent t-tests of equal variance were used to compare the change in SBP and DBP from pre-exercise to within 1-minute post-exercise (continuous, mmHg) between groups (categorical, two levels: stroke, non-stroke).

Both protocols were matched for relative target intensity; however, it was possible that perceived intensity may have differed between stroke and non-stroke groups due to individual perceptual differences. Additionally, the magnitude of the post-exercise BP response is intensity dependent.^26,41,42^ Therefore, an independent t-test of equal variance was used to compare rating of perceived exertion (continuous, n/10) between groups (categorical, 2 levels: stroke, non-stroke).

In the adjusted analysis, we performed a two-way analysis of covariance (ANCOVA) to examine the effect of stroke status (categorical, two levels: stroke, non-stroke), baseline SBP and DBP (continuous, mmHg), and activity level (continuous, minutes/week) on change in SBP and DBP from pre-exercise to within 1 minute post-exercise (continuous, mmHg).

Significance for adjusted and unadjusted analyses was set at 0.05. All statistical analyses were conducted using Stata 19 (StataCorp, College Station, Texas, United States).

## RESULTS

### Participants

In the stroke group, 15 participants were enrolled in the main trial, but data for 4 participants were not included due to absence on the designated training session (n=2) and brachial BP measurement error (n=2). Thus, the final sample included 11 participants (6 males, 5 females), aged 57.3 ± 10.5 in the stroke group. For the non-stroke group, data on all 19 participants (8 males, 11 females), aged 22.8 ± 4.0 were available. Table 1 displays participant characteristics by group. Perceived intensity from the single 8-repetition set of horizontal leg press was similar between groups (Mean difference (MD) = 0.05, 95%CI: -1.10 to 1.20, (t(28)=0.09, p=0.93).

**Figure 1.**
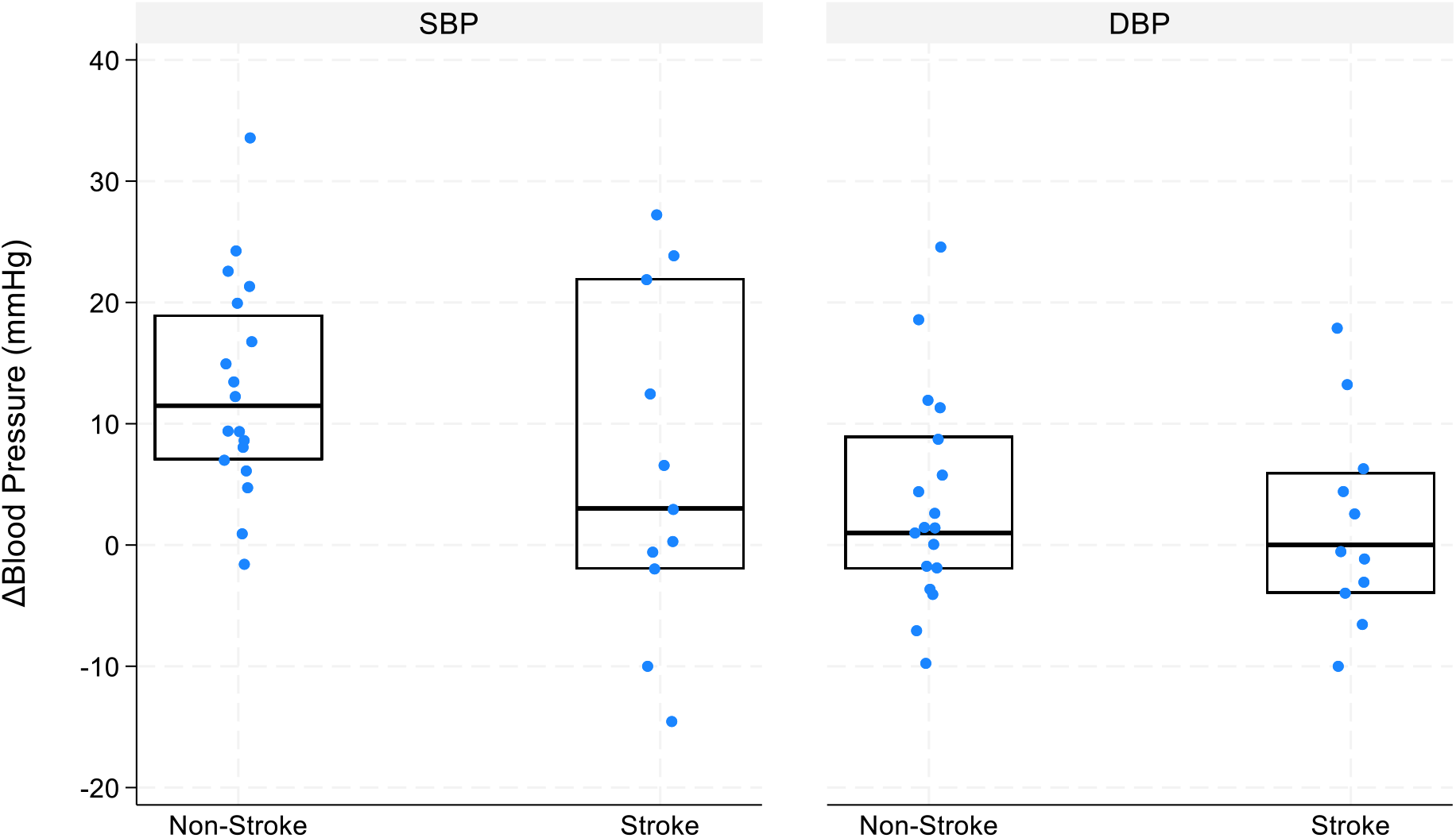
Systolic and diastolic blood pressure change post-exercise by stroke status. Median change in blood pressure represented with center line, and interquartile range (25^th^-75th percentiles) represented with box. Tails represent minimum and maximum points.

**Table 1.**
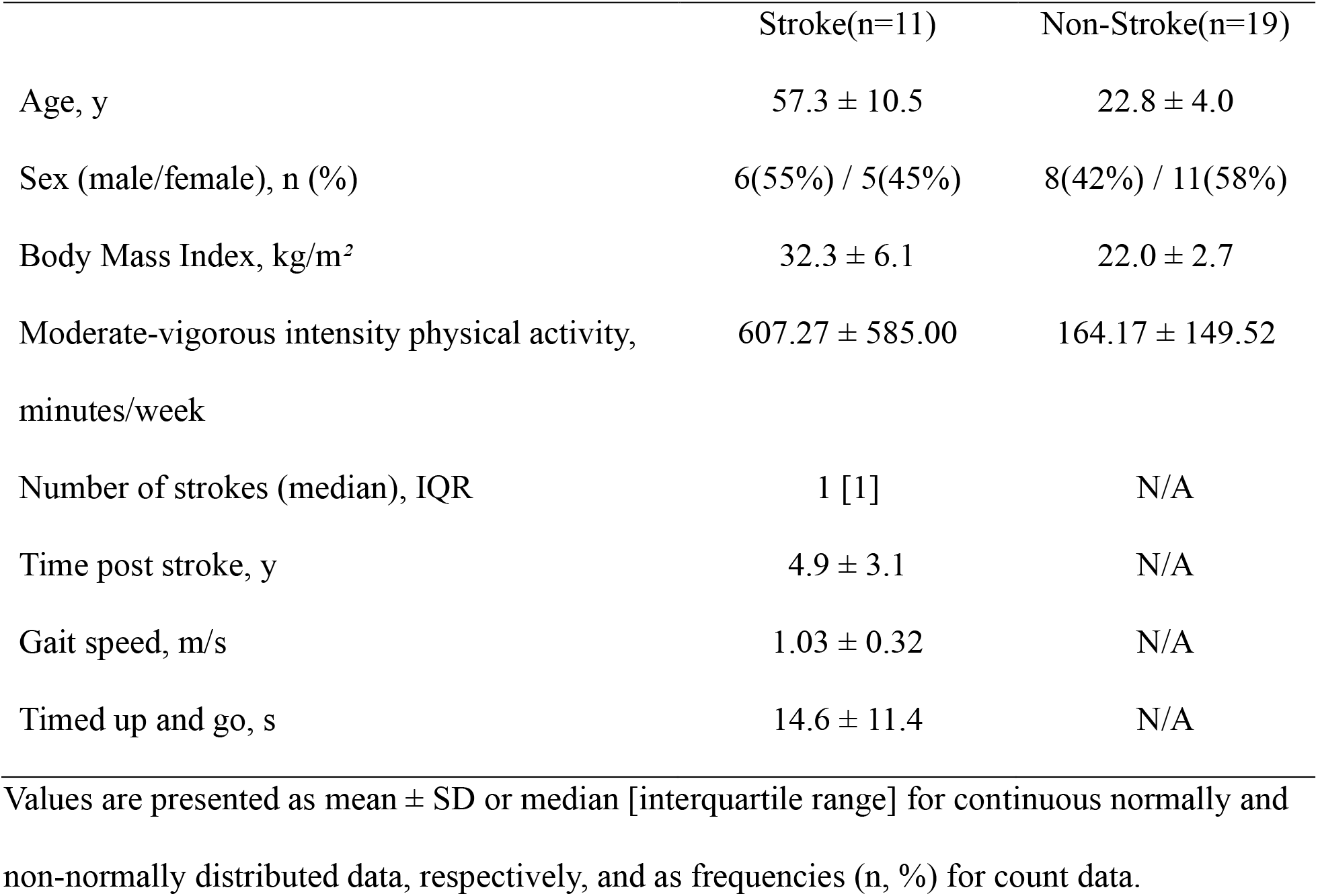
Participant characteristics by group.

|  | Stroke(n=11) | Non-Stroke(n=19) |
| --- | --- | --- |
| Age, y | 57.3 ± 10.5 | 22.8 ± 4.0 |
| Sex (male/female), n (%) | 6(55%) / 5(45%) | 8(42%) / 11(58%) |
| Body Mass Index, kg/m<sup>2</sup> | 32.3 ± 6.1 | 22.0 ± 2.7 |
| Moderate-vigorous intensity physical activity,<br>minutes/week | 607.27 ± 585.00 | 164.17 ± 149.52 |
| Number of strokes (median), IQR | 1 [1] | N/A |
| Time post stroke, y | 4.9 ± 3.1 | N/A |
| Gait speed, m/s | 1.03 ± 0.32 | N/A |
| Timed up and go, s | 14.6 ± 11.4 | N/A |

Values are presented as mean ± SD or median [interquartile range] for continuous normally and non-normally distributed data, respectively, and as frequencies (n, %) for count data.

### Blood Pressure Response

Pre-exercise SBP and DBP were 129.0±16.0 mmHg and 77.1±8.0 mmHg in the stroke group, and 116.3±13.4 mmHg and 71.7±5.6 mmHg in the non-stroke group. Table 2 displays mean pre-exercise and exercise responses by group.

**Table 2.** Mean exercise response by group.

|  | Stroke(n=11) | Non-Stroke(n=19) | p-value |
| --- | --- | --- | --- |
| Pre-exercise |  |  |  |
| Systolic Blood Pressure, mmHg | 129.0 ± 16.0 | 116.3 ± 13.4 | 0.03 |
| Diastolic Blood Pressure, mmHg | 77.1 ± 8.0 | 71.7 ± 5.6 | 0.06 |
| Post-exercise change |  |  |  |
| Systolic Blood Pressure, mmHg | 6.18 ± 13.83 | 9.79 ± 16.24 | 0.54 |
| Diastolic Blood Pressure, mmHg | 1.72 ± 8.37 | 7.11 ± 17.37 | 0.35 |
| Rating of Perceived Exertion, n/10 | 4.77 ± 0.90 | 4.82 ± 1.73 | 0.93 |

Values are presented as mean ± SD.

In the unadjusted analysis, there were no differences between groups in changes in SBP (MD = 3.61, 95%CI: -8.36 to 15.57, t(28)=0.62, p=0.54) or DBP (MD = 5.38, 95%CI: -6.11 to 16.86, t(28)=0.96, p=0.35) (Figure). After controlling for baseline SBP, baseline DBP and physical activity levels, there were still no group differences in change in SBP (stroke: marginal mean = 10.07, 95%CI: 1.65 to 18.50; non-stroke: marginal mean = 10.57, 95%CI: 4.48 to 16.66; F(3,26)=6.81, p=0.08; Table 3) or DBP (stroke: marginal mean = 3.26, 95%CI: -2.55 to 9.06; non-stroke: marginal mean = 2.68, 95%CI: -1.68 to 7.03; F(3,26)=3.96, p=0.27; Table 4).

**Table 3.**
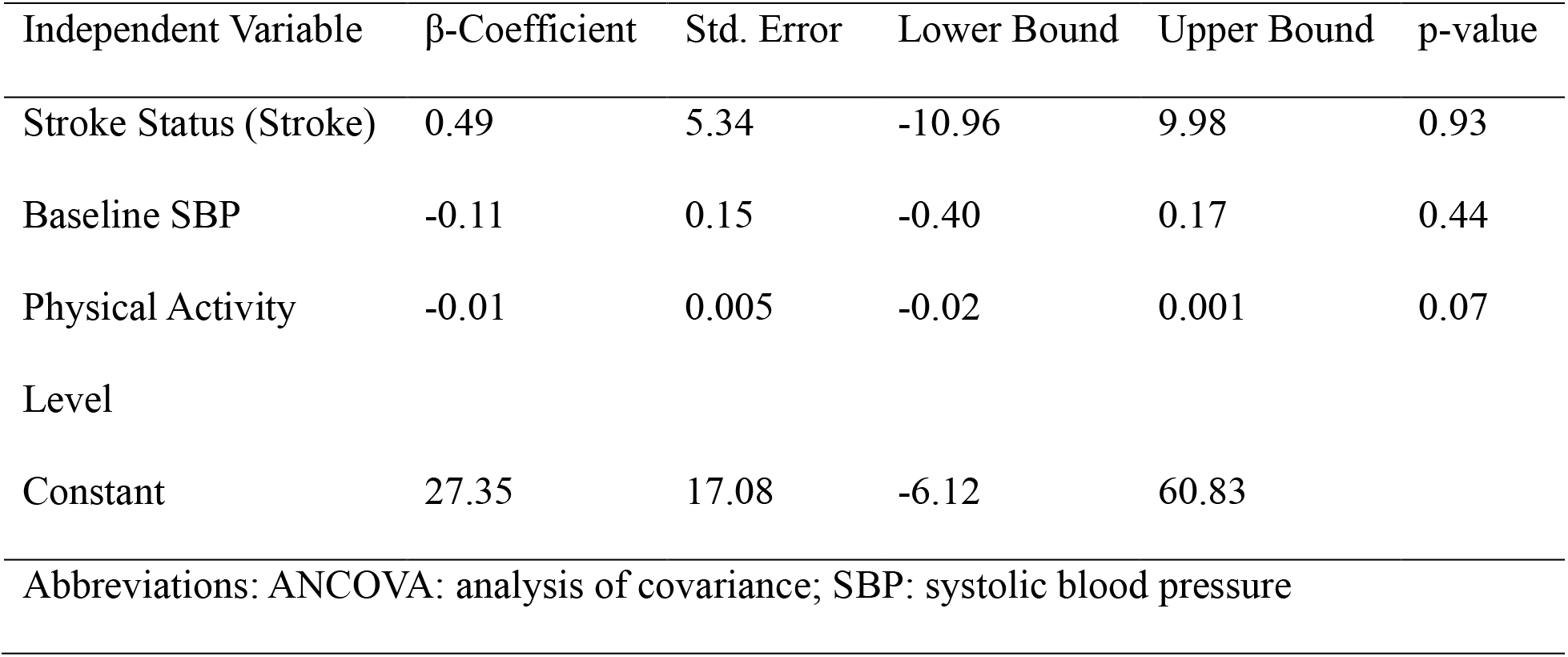
Adjusted analysis of two-way ANCOVA on SBP response immediately following exercise.

| Independent Variable | β-Coefficient | Std. Error | Lower Bound | Upper Bound | p-value |
| --- | --- | --- | --- | --- | --- |
| Stroke Status (Stroke) | 0.49 | 5.34 | -10.96 | 9.98 | 0.93 |
| Baseline SBP | -0.11 | 0.15 | -0.40 | 0.17 | 0.44 |
| Physical Activity Level | -0.01 | 0.005 | -0.02 | 0.001 | 0.07 |
| Constant | 27.35 | 17.08 | -6.12 | 60.83 |  |

Abbreviations: ANCOVA: analysis of covariance; SBP: systolic blood pressure

**Table 4.**
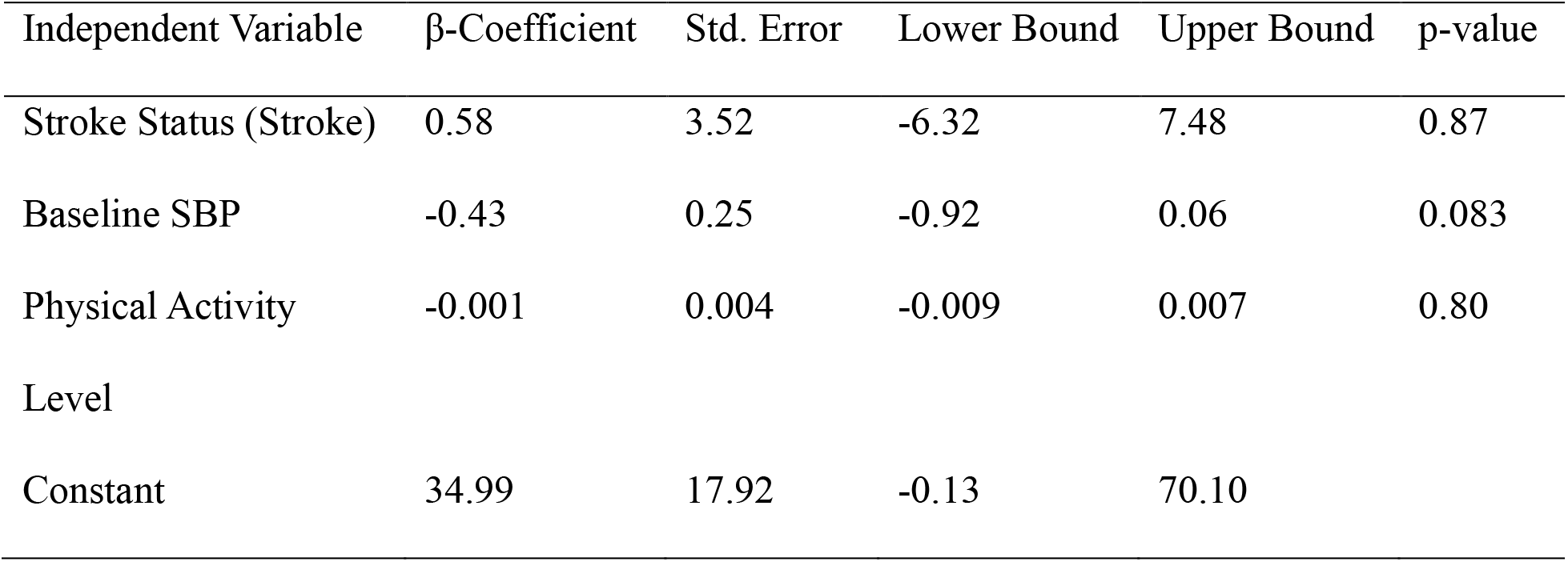
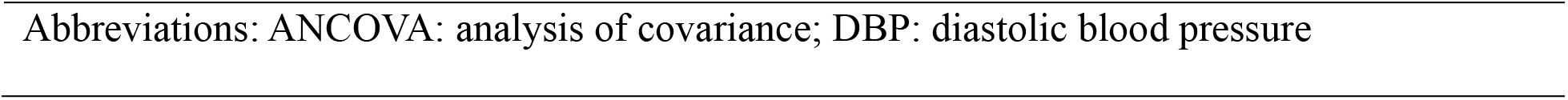
Adjusted analysis of two-way ANCOVA on DBP response immediately following exercise.

| Independent Variable | β-Coefficient | Std. Error | Lower Bound | Upper Bound | p-value |
| --- | --- | --- | --- | --- | --- |
| Stroke Status (Stroke) | 0.58 | 3.52 | -6.32 | 7.48 | 0.87 |
| Baseline SBP | -0.43 | 0.25 | -0.92 | 0.06 | 0.083 |
| Physical Activity Level | -0.001 | 0.004 | -0.009 | 0.007 | 0.80 |
| Constant | 34.99 | 17.92 | -0.13 | 70.10 |  |

Abbreviations: ANCOVA: analysis of covariance; DBP: diastolic blood pressure

## DISCUSSION

Our pragmatic research approach examined the BP response to a single acute bout of resistance exercise in individuals with and without stroke. We hypothesized that the stroke group would demonstrate a greater reduction in SBP and DBP following acute resistance exercise compared to the non-stroke group; however, our results indicate that there were no significant between-group differences. Instead, both groups exhibited an increase in SBP and DBP immediately following resistance exercise. While these findings may appear unexpected, it is possible that sample characteristics and methodological factors may help explain the observed responses.

These findings are inconsistent with previous literature that have reported greater magnitude reductions in BP in populations with autonomic dysfunction (i.e., hypertension) following resistance exercise.^29,30^ However, the extent to which these prior findings are applicable to individuals in the chronic phase of stroke recovery remains unclear. Although BP regulation is often impaired early after stroke,^13–15^ there is limited evidence examining these trends during the chronic phases of stroke recovery. Further, the participants in the current study were between 1 and 9 years post-stroke, representing a sample with potentially substantial variability in residual autonomic impairment. Consequently, post-exercise BP responses may not be uniform across our sample comprised of individuals at varying times after stroke, and it is therefore possible that the absence of differential BP responses between groups may reflect in part the heterogeneous nature of chronic stroke recovery.

Despite adjustment for resting hemodynamics and physical activity levels, the groups were not age matched. This is important as BP regulation worsens with older age,^43^ contributing to our prior hypotheses wherein we expected greater reductions in BP following acute resistance exercise in the older stroke group. While these findings are not matched in the current study, it is possible that this may be owed to the stroke group participating roughly 10 times more minutes of moderate-to-vigorous intensity physical activity per week than typical stroke populations (607 compared to 61).^44^ Exercise training has a beneficial effect on endothelial function,^45,46^ baroreflex sensitivity,^47^ and neuroplastic changes within autonomic cardiovascular control pathways.^48^ Although, age-related impairments in BP regulation including arterial stiffness and autonomic dysfunction,^49^ paired with stroke-related impairments including increased muscle sympathetic nerve activity^50^ and decreased baroreflex sensitivity,^51,52^ would be expected to exacerbate the decrease in BP following exercise, these effects may have been attenuated by the vascular and neurological adaptations associated with regular exercise training. Therefore, these results may not be applicable to individuals with lower activity levels after stroke and more work is needed to elucidate the specific mechanisms underlying these potential chronic adaptations in samples that better represent chronic stroke populations.

Methodological factors may have similarly contributed to the lack of differential responses between groups and may also help explain the increased BP response that is inconsistent with the post-exercise hypotension literature. The intensity of exercise plays a crucial role in the magnitude and duration of the post exercise hypotension response,^41,42^ largely due to a more pronounced reduction in peripheral vascular resistance.^53^ This intensity dependent relationship has also been replicated in previous stroke literature,^26^ reinforcing the relevance of intensity matching. In the current study, target intensity was matched (7/10 modified Borg Scale ≈ 70% 1-

RM).^35,36^ Although perceived level of effort was lower than the prescribed exercise intensity (<5/10 compared to 7/10) in the stroke group, potentially reflecting the cautious approach taken by exercise trainers to progress participants to higher workloads, the successive nature of this secondary analysis allowed for the alignment of perceived levels of effort between groups. Therefore, any observed differences in BP responses to resistance exercise are unlikely to be attributable to disparities in exercise intensity. However, given that low- to moderate-intensity exercise typically elicits weakened decreases in the post-exercise BP response and the perceived intensity of exercise in the current studies marginally exceed the threshold set at 40% of 1 repetition maximum required (∼4/10 rating of perceived exertion)^36^ to elicit post-exercise hypotensive response,^19^ it remains possible that the resistance exercise stimulus may not have been vigorous enough to differentiate differences in BP recovery between our stroke and non-stroke samples. As such, we recommend future studies to consider the use of more intense exercise to examine potential differences following high-intensity resistance exercise in this population.

Moreover, the rate of BP decline after exercise is said to be the largest immediately following exercise for up to 30 seconds;^33,34^ however, the stroke literature suggests delayed cardiovascular recovery following intense exercise.^24^ As such, we measured BP within the first 60 seconds following the exercise set but acknowledge it is possible that we may have missed individual differences in BP recovery due to stroke induced delays in post-exercise BP recovery. Post-exercise BP was also measured as a discrete single measurement to reflect common practice in community exercise programs, and the BP response to acute resistance exercise was calculated as the difference between pre- and post-exercise measurements. However, we were thus unable to capture potential differences in BP during exercise and the rate of BP decline thereafter that could be afforded by continuous beat-to-beat^33^ or longer post-exercise duration measurements.^34^

## Data Availability

All data produced in the present study are available upon reasonable request to the authors

## Limitations

This study was not stastically powered nor was it meant to provide an explanation into physiological mechanisms, but rather, was designed to provide practical and preliminary insights into differences that may be associated with the safety of resistance exercise in individuals with stroke. Consequently, the confidence intervals for both groups were wide and potentially included meaningful between-group differences that may differentiate BP responses in individuals with and without stroke. The small sample size also precluded the statistical analysis of other explanatory variables including antihypertension medication (stroke group n=5, 45%), which may have suppressed the suspected rise in BP during exercise and therefore may have influenced the subsequent BP measurements following exercise. We also acknowledge that slight postural differences in the leg press exercise (90-degree horizontal leg press in stroke, 45-degree semi-recumbent leg press in non-stroke) may have affected BP regulation during and after exercise,^54^ due to changes in the hydrostatic gradient and the subsequent baroreflex response between different seated positions.^55^

## Conclusion

The pragmatic nature of this study provides insight into the emerging field of cardiovascular responses to resistance exercise in people with stroke. Overall, there were no differences in changes in SBP and DBP following resistance exercise between people with and without stroke, even after controlling for baseline BP and physical activity levels; however, more work is needed to support these findings. This study lays the foundation for future work to elucidate mechanisms underlying BP regulation following acute resistance exercise to equip healthcare professionals working with stroke with the necessary evidence to inform safe exercise prescription.

